# “Spectromics”: Holistic Optical Assessment of Human Cartilage via Complementary Vibrational Spectroscopy for Osteoarthritis Diagnosis

**DOI:** 10.1101/2023.12.21.23300367

**Authors:** Hiroki Cook, Anna Crisford, Konstantinos Bourdakos, Douglas Dunlop, Richard OC Oreffo, Sumeet Mahajan

## Abstract

Osteoarthritis (OA) is the most common degenerative joint disease, presented as wearing down of articular cartilage and resulting in pain and limited mobility for 1 in 10 adults in the UK. ^1^ There is an unmet need for patient friendly paradigms for clinical assessment that do not require ionising radiation (CT), exogenous contrast enhancing dyes (MRI), biopsy, and/or instrumentation approaches (arthroscopy or endoscopy). Hence, techniques that use non-destructive, near- and shortwave infrared light (NIR, SWIR) may be ideal providing for non-invasive, label-free and deep tissue interrogation. This study demonstrates multimodal “spectromics”, low-level abstraction data fusion of non-destructive NIR Raman scattering spectroscopy and NIR-SWIR absorption spectroscopy, providing an enhanced, interpretable “fingerprint” for diagnosis of OA in human cartilage. Samples were excised from femoral heads post hip arthroplasty from OA patients (n=13) and age-matched control (osteoporosis) patients (n=14). Under multivariate statistical analysis and supervised machine learning, tissue was classified to high precision: 100% segregation of tissue classes, and a classification accuracy of 95% (control) and 80% (OA), using the combined vibrational data. There was a marked performance improvement (5 to 6-fold for multivariate analysis) using the spectromics fingerprint compared to results obtained from solely Raman or NIR-SWIR data. Furthermore, discriminatory spectral features in the enhanced fingerprint elucidated clinically relevant tissue components (OA biomarkers). In summary, spectromics provides comprehensive information for early OA detection and disease stratification, imperative for effective intervention in treating the degenerative onset disease for an aging demographic.

## Introduction

Osteoarthritis (OA) presents a major public health challenge recognised as a serious burden for the individuals affected, healthcare systems, and resulting in significant national and global socioeconomic costs.^1,2^ Current modes of assessment of articular cartilage in the clinic are typically invasive (endoscopy, arthroscopy), destructive (biopsy, histochemistry), incorporating ionising radiation and/or exogenous contrast (CT, X-ray, MRI).^3,4^ Each modality provides different levels of qualitative morphological information of the tissue to assess the health of the patient but typically necessitate clinical evaluation and treatment prognosis. Critically, none of these indicated techniques provide a definitive diagnosis for OA, which needs to be confirmed by secondary methods.

Current gold standard approaches for diagnosing OA rely on the detection of pain, morphological changes (joint space narrowing, osteophytes formation), or accumulation of synovial fluids.^5,6^ OA presents degradation and loss of articular cartilage, the lubricating and shock absorbing inter-joint layer, developed over many years, and which can pre-date symptoms over decades.^7^ Hence, early diagnosis is crucial for effective and timely intervention to reduce pain, improve mobility, and patient quality of life. To date, there is no cure for OA, rather treatments are focused on alleviating inflammatory symptoms or interventional surgery including arthroplasty and prosthetic joint implant.^8–10^ Detection of pre-pathomorphological and biochemical changes will inform new and earlier forms of pharmacological and lifestyle interventions to alter the course and progression of the disease.^11,12^

Near- and Shortwave Infrared (NIR, SWIR) absorption and spontaneous Raman scattering are highly sensitive to structural and biochemical changes offering an innovative approach for early detection associated with the onset of OA, using spectral biomarkers. This can be ideal as both Raman and NIR-SWIR absorption spectroscopy can be carried out with minimal sample preparation, conducive to native *in situ* tissue assessment. Moreover, both NIR-SWIR absorption and Raman spectroscopies can utilise the biological ‘optical’ transparency windows. Such windows exist in various native human and animal tissue types and are quantified by the absorptive and scattering effect of common endogenous chromophores. In these regions, non-ionising optical light undergoes reduced scattering and absorption, facilitating deep penetration.^13–15^ Depth penetrations in AC tissue are recorded up to 5 mm, indicating high suitability for optical assessment.^16,17^ Since the normal thickness of human cartilage is 1 – 3 mm, optical interrogation in the spectral range of 1.4 – 2.5 µm is considered optimal for AC tissue assessment.^17,18^ Thus, spectroscopic techniques such as NIR-SWIR absorption and Raman spectroscopy (if carried out with NIR or SWIR excitation) offer valuable chemometric and structural information at depth for non-destructive, non-invasive *in vivo* clinical assessment.

NIR-SWIR absorption spectroscopy has been shown to be sensitive to structural and compositional changes resulting from loss or alteration of the tissue extracellular matrix (ECM).^19^ The spectral response provides information relevant to structural and functional characteristics, important in the assessment of degeneration of cartilage.^20–23^ Specifically, absorbance bands in the NIR and SWIR are overtones and combinations of the fundamental vibrations of O-H, C-H, N-H, and S-H bonds which form the molecular framework of the tissue. ^16,19,24^ As such, NIR-SWIR absorption spectroscopy can offer a non-destructive method to determine thickness, biomechanical properties, and composition of articular cartilage especially water fractions for evaluation, prediction, and monitoring of OA progression. ^17,18,21,22,25,26^

Spontaneous Raman scattering spectroscopy, is insensitive to water, and the spectral region between 1800 - 800 cm^-1^ is particularly sensitive to structural and skeletal vibration modes, thus ideal for biological tissue characterisation.^27^ Raman spectroscopy using NIR excitation has been employed by a number of groups in the diagnosis of osteoarthritis, with biochemical and biomechanical change in human and preclinical models correlating with gold standard assays. ^18,25,28^ Spectral features including those associated with collagen, GAG, and PG (major proteins of the ECM), water fraction, lipid and amides have proved efficacious as preclinical and prepathomorphological biomarkers.^29–31^ Specific biochemical distributions have been mapped up to depths of 0.5 mm with NIR excitations ^27,32–34^ and OA relevant signals (depth and GAG) collected at depths >10 mm under spatially offset Raman spectroscopy (SORS) geometries.^35^

NIR-SWIR absorption and NIR-excited Raman spectroscopy have been used to interrogate cartilage and shown potential for OA diagnosis, especially through evaluation of depth-dependent features.^36^ Since these techniques are mediated by different optical phenomena, namely changes in dipole moment (overtones of vibrational modes) and changes in polarization, respectively, they offer complementary information. As such, these modalities can be combined to yield a more holistic chemical ‘fingerprint’ of the sample of interest. This study presents the first demonstration of an elegant combination of vibrational spectroscopy techniques operating in the transparency windows to augment their diagnostic assessment potential of human articular cartilage. Raman scattering and NIR-SWIR absorption signatures are concatenated through low-level abstraction data fusion, into a new “spectromic” fingerprint. ^37–39^ The fused data was subjected to statistical and machine learning analysis, namely supervised Principal Component Analysis – Linear Discriminant Analysis (PCA-LDA) and supervised support vector machine (SVM).^37^ The new fingerprint facilitated improved classification accuracy and delineation between control and osteoarthritic AC tissue compared to spectra from each technique individually. The low-level abstracted data is directly interpretable since significant spectral features used to classify the tissue can highlight clinically relevant biomarkers, and is compatible with various statistical and machine learning assessments. The current studies demonstrate the efficacy and power of a spectromics approach in its ability to provide a holistic assessment of human cartilage tissue for OA diagnosis, with therapeutic implications for an increasing aging population.

## Experimental

### Cartilage Samples and Preparation

Articular cartilage samples, obtained with full ethical approval and patient consent (REC reference 18/NW/0231), were excised from human femoral heads manually using a scalpel blade. Cartilage tissue slices were taken parallel to the femoral head surface, as deep as the subchondral bone. Cartilage slices were fixed in 4% paraformaldehyde (PFA) for 72 hrs and stored, refrigerated, in phosphate buffered saline.

Tissue storage conditions were found to be compatible with vibrational spectroscopy characterisation, with consistent Raman spectral responses reproduced despite storage periods approaching 6 months.

For spectroscopic and imaging analysis, AC samples were cut into square slices, side lengths of the order of 10’s of mm, thickness of ∼1 mm. For each sample, the “superficial surface” describes the outermost layer of cartilage on the femoral head (in contact with the acetabular cup) and “deep side” the layer proximal to the subchondral bone. This terminology reflects the zonal stratified structure reported for cartilage wherein the superficial-, middle-, and deep zones (SZ, MZ, DZ) and calcified zones contain varying collagen fibre orientation and composition.^4,10,29,33^

Samples obtained from osteoarthritic (OA) femoral heads serve as the diseased AC model, while samples from osteoporotic (OP) femoral heads served as the control model. Both classes of tissue were obtained as consequence of interventional arthroplasty surgery. Analysis of human samples enabled paradigm development to inform and model clinical application. Patient information included only patient age and gender, and classification between OA and OP. Classification was confirmed by the consultant orthopaedic surgeon with OA samples typically Grade 3 & 4 (late stage) OA progression. Lack of access to healthy mature human cartilage necessitated the use of AC from OP femoral heads as the “healthy” control model tissue. OA pathology results in severe thinning of AC around the femoral head such that the subchondral bone is typically exposed in large regions. OA samples were taken from areas where AC remained present.

Samples were selected in order to match and be equally distributed across anthropometric parameters. Spectra were collected from AC of n=13 OA patients and n=14 OP patients for this proof-of-concept study.

### Raman Scattering Microspectroscopy

Raman spectroscopy of the cartilage samples was carried out *via* modification of a previously reported protocol. ^36,41^ Samples of AC were placed on a quartz slide, “superficial” side up, and their spectra centred at 1200 cm^-1^ (614 – 1722 cm^-1^) captured in reflectance geometry. Measurements were carried out on a Renishaw InVia microscope system with samples excited using a 785 nm laser focused through a Leica 50x (0.75 NA) short working distance (∼200 µm) objective. Renishaw WiRE 4.1 software was used to collect data and set measuring parameters. The system was calibrated to the 520 cm^-1^ peak of a silicon standard before each experiment and cosmic rays removed after acquisition. Background-subtraction was carried out by subtraction of the spectrum of a blank quartz slide, taken with the same acquisition parameters as experimental samples. Spectral resolution was recorded as Δλ ∼1.1 cm^-1^.

Spectral mapping of each sample was achieved by movements of the sample stage in random steps of the order of 100’s µm to measure across the tissue surface. For each position, a mean average spectrum of 3 acquisitions with exposure time of 5 seconds was recorded. A modal average of 3 samples were investigated for each patient with 10 spectra measured for each sample.

### NIR-SWIR Absorption Spectroscopy

NIR-SWIR spectroscopy of the cartilage samples was carried out on a homemade benchtop system. Samples of AC were placed on a gold-coated mirror slide, “superficial” side up, and spectra between 11,127 – 3993 cm^-1^ (899 – 2504 nm) captured in transreflectance geometry. Incident excitation light was provided by a broadband halogen lamp (HL-2000-FHSA-LL, Ocean Insight) emitting as a blackbody across the NIR-SWIR range, and signal collected via an OceanOptics NIR Quest 2.5+ spectrometer. Both were coupled to a ferrule fibre optic reflectance probe with a profile of 6 annular fibres for excitation and 1 central fibre for collection. ^22^ Two planoconvex uncoated lenses collimated and focused light onto sample, allowing for contact-less measurements. Spectral resolution of the spectrometer was quoted by the manufacturer at Δλ ∼6.3 nm.

Spectral mapping of each sample was achieved by manual sample scanning in random steps of the order of 100’s um across the surface of the tissue. For each position, a mean average spectrum of 100 acquisitions with exposure time of 10 milliseconds was recorded. A modal average of 3 random samples were investigated for each patient and 10 spectra measured for each sample.

### Spectral Pre-processing

Spectra were labelled with the patient’s age, sex, and OA/OP classification. The modal average was 30 spectra per patient (10 spectra per sample, 3 samples per patient). Spectral data underwent pre-processing transformations prior to classification via multivariate analysis, carried out in iRootLab (0.15.07.09-v) toolbox within MATLAB R2020a software (MathWorks).^42^

Raman scattering spectra were treated with 5^th^-order polynomial baseline correction, to eliminate slow varying offset attributed to interference of fluorescence and Mie scattering.^41,43,44^ Wavelet de-noising via 6 level Haar wavelet thresholding minimised random spectral noise without affecting signal quality.^42,45^ Rubber-banding baseline correction anchored the primary and terminal ends of each spectra to the horizontal axis, before vector normalisation. ^41,46 47^

NIR-SWIR absorbance spectra were first treated with a 1^st^-derivative transformation to elucidate subtle features, as well as eliminate baseline offset, linear trends, and interference from light scattering. ^16,44^ To mitigate subsequently increased noise (reduced SNR), a 2^nd^ order Savitsky-Golay smoothing filter was applied (suitable for vibrational spectroscopy data) with 9 filter coefficients (reducing noise whilst preserving information). ^16,44,47^ A rubber-banding baseline correction was also applied, suitable for later concatenation since the terminal value of the Raman region could marry with the starting value of NIR-SWIR. Finally, a further application of a 6 level, Haar wavelet de-noising transformation before vector normalisation served to make Raman and NIR-SWIR spectral peaks a compatible magnitude.

### Concatenation

The spectromics spectral fingerprint was built by concatenation of data from the mean average pre-processed Raman spectra to the mean average pre-processed NIR-SWIR data from a given patient. The mean spectra recorded from each of the techniques was analysed given the spatial locations were not correlated between Raman and NIR-SWIR spectroscopy measurements. This also ensured that the spectra were truly representative of the whole sample for each patient. Concatenation is thus an abstraction of the data wherein the overall spectral shape is preserved (Raman scattering and NIR-SWIR absorbance) and the independent variable becomes a reference point in the spectromics fingerprint. Previously we have reported on the low level fusion between two Raman spectra excited at different wavelengths for accurate, label-free characterisation of bacterial pathogens.^45,48^ This concatenation process resulted in data fusion with Raman spectra accounting for the first 1011 independent variables (data points) and NIR-SWIR the last 512 points. ^37–39^

### Multivariate Analysis

#### PCA-LDA

Each of the pre-processed Raman, NIR-SWIR, and spectromics data set was mean centred before Principal Component Analysis (PCA).^16^ PCA scores represented variance in the sample direction and highlighted clustering patterns related to chemical similarities/dissimilarities between samples. Loadings describe the signal variance across the independent variable for identifying spectral regions with high degree of significance to the PCA scores distribution.^44^ PCA could not systematically classify samples alone and instead required further classification techniques.^49^ Herein the first 10 and first 20 PC’s (responsible for the majority of variance in the dataset) were selected for supervised classification via Linear Discriminant Analysis (LDA). PCA-LDA assigns the cartilage samples to their predicted groups, “Control” and “OA” cartilage.^50^ This algorithm calculated the Mahalanobis distance between samples for each class as a measure of tissue class segregation. ^44^ Classification for diagnosis here was supervised, labelled *a priori* via gold standard assessment from the orthopaedic surgeon. Though LDA is a parametric method and assumes samples hold a normal distribution, it was considered robust enough for spectroscopic data, and by applying to the foremost PCA scores maintains that the number of spectral variables was larger than the number of samples.^44^

#### SVM

Support Vector Machine (SVM), a supervised binary linear machine-learning classifier, was applied to each the Raman, NIR-SWIR, and spectromic fingerprints to quantify classification accuracy. A k-fold cross validation method was used to train the Gaussian classifier to model “Control *vs* OA Cartilage”, with the optimal values for the parameters for *c* and *γ* determined via a grid search function. ^42,50^ This, was carried out for k=3 and leave-one-out cross validation to determine training and test sub-datasets. Confusion matrices of each SVM model, built on the Raman, NIR-SWIR and spectromics fingerprints, describe the rate of correct group assignation when applying the trained model to the test dataset.

#### Quality Parameters

Quality parameters were calculated using the accumulative hits of the classification model, describing number of true positives (TP), true negatives (TN), false positives (FP), and false negatives (FN). This quantified the classification accuracy of the respective models (Raman, NIR-SWIR, spectromics) on a test dataset (no *a priori* indication). ^44^ The comparator metrics, equations, and relevance to the technique are summarised in Table 1.

**Table 1:**
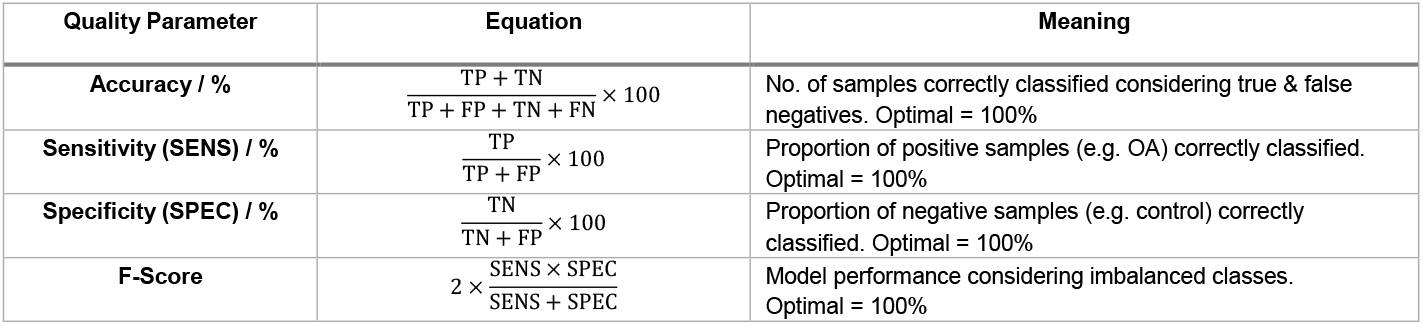
Quality parameters used to evaluate model classification performance. Here TP means True Positive, FP false positives, TN true negatives, and FN false negatives. Adapted from Medeiros-De-morais et al.^44^.

#### Feature Extraction: Spectral Biomarkers

Feature extraction was performed on the spectromics data set to ascertain potential spectral biomarkers that account for the biggest differences between the OA and control tissue groups. The key biomarkers were identified by observing agreement between the highest weighted results of several independent statistical tests. Each process was carried out within IRootLab. ^42^

PCA-LDA Loadings allowed feature extraction by first identifying the 3 highest absolute loading coefficients of the first LD component corresponding to the highest contributing PC’s. The loadings of the top 3 discriminant PC’s were in turn plotted to identifying the highest absolute peaks in the wavenumber direction. Cluster Vector Analysis following PCA-LDA highlighted PC’s that represent the best samples’ clustering and their loading vectors were combined.^42,44^ Both approaches highlighted the most pertinent spectral regions for tissue classification in the PCA-LDA model, and the top 20 wavenumbers were recorded for each.

Differences Between Mean Spectra (DBMS) identified biochemical alterations between the mean spectra of the control AC tissue (reference) and of the OA tissue (investigated sample). The top 20 most distinguishing features were recorded.

The Student’s T-Test and Mann-Whitney U-Test were each applied to the spectromics fingerprint to test the probability of correct classification. The former assumed a normal (Gaussian) distribution and the latter non-parametric (arguably with less bias), both carried out for completeness.^42,44^ The -log10 of the P-value for the T- and U-Test for each wavenumber was plotted. Feature extraction was carried out by identifying the 20 largest peaks above a threshold of p=0.01 (99% confidence interval).^44^

Finally, Feature Forward Selection was carried out via standard protocol in the iRootLab toolbox. Here a binary classification was iterated to find the optimal features for class segregation, trained using a random 90% portion of the data, tested on the remaining 10%. Feature histograms counted how many times particular wavelength were selected, and displayed the most important features for distinguishing between the condition (OA) and reference class (control).^51,42^ The 7 most segregating peaks were recorded from the FFS histogram.

Candidate biomarker spectral features from the concatenated fingerprint were mathematically translated to the corresponding Raman and NIR-SWIR spectra for assignment.^44^ NIR-SWIR features were corroborated through comparison to the nearest 1^st^ derivative and corresponding 2^nd^ derivative transformed spectral peaks. This allowed correct attribution to the zero-order NIR-SWIR spectra.

## Results and discussion

### Deeply penetrating vibrational spectroscopy of articular cartilage

Representative spectra obtained with back-scattered Raman scattering and transreflected SWIR absorption spectroscopy of articular cartilage are shown in Fig 1. The class means for control and OA tissue from Raman spectra and the corresponding 1^st^ order derivative NIR-SWIR spectra for the same patient, used to form the spectromics concatenated fingerprint are shown. For NIR-SWIR absorption spectra it was found that transreflectance geometry measurements (non-contact, highly reflective substrate) produced the same spectral response as for transmission geometry measurements (illumination and collection from either side of the sample), recorded in-house, and for backscattered fibre probe measurements (in contact, diffuse reflectance), recorded in literature.^16,22 21,22^ However, transreflectance measurements necessitate the use of a highly reflective substrate. Native *in vivo* tissue classification will require diffuse reflectance measurements (subchondral bone as natural substrate), nevertheless, NIR-SWIR results presented here enable us to provide proof-of-concept.

**Fig. 1:**
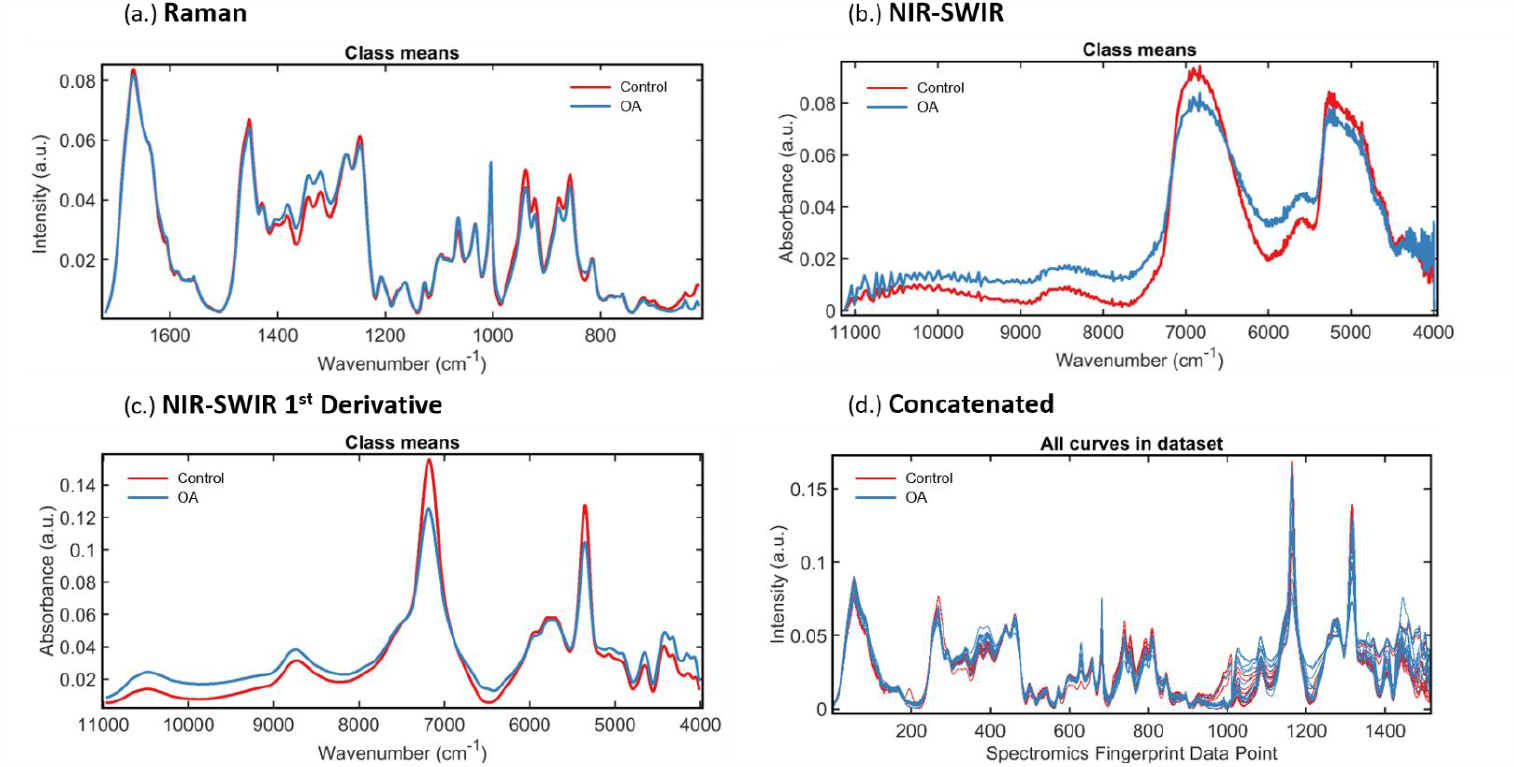
Typical vibrational spectroscopy fingerprints of articular cartilage, (a.) Represents Raman spectroscopy (614 − 1722 cm^-1^) pre-processed to correct background signal and random noise, and vector normalised, (b.) i. Represents typical NIR-SWIR absorption (11,127 − 3993 cm^-1^) minimally pre-processed to remove extraneous signal, and normalised, (c.) Represents NIR-SWIR spectra with 1st Derivative pre-treatment to elucidate subtle peaks, smoothed and normalised, (d.) Concatenated Spectromics Fingerprint showing abstracted Raman spectra (1011 data points) fused to NIR-SWIR spectra (512 data points)

### Multivariate Analysis Modelling: Improved Tissue Classification using Spectromics

Raman scattering spectra, NIR-SWIR absorption spectra, and Concatenated spectra were assessed under multivariate statistical analysis to determine tissue classification accuracy. The data represented a mean average spectra characteristic for each of n=13 osteoarthritis and n=14 control model patients.

Classification via PCA clustering alone proved inconclusive for each spectral modality since significant overlap existed between groupings of control and OA spectra. This could be due to the relatively small number of patient samples, exacerbated by the lack of spatially correlated Raman and NIR-SWIR data. LDA based on the first 10 PCs and the first on 20 PCs showed inter-group differences, measured in Mahanobolis distance between control (negative class) and OA cartilage (positive), displayed in Fig. 2. Quality parameters were calculated from the corresponding proportion of TP, FN, TN, and FP segregation of samples, summarised in Table 2. ^44^

**Table 2:**
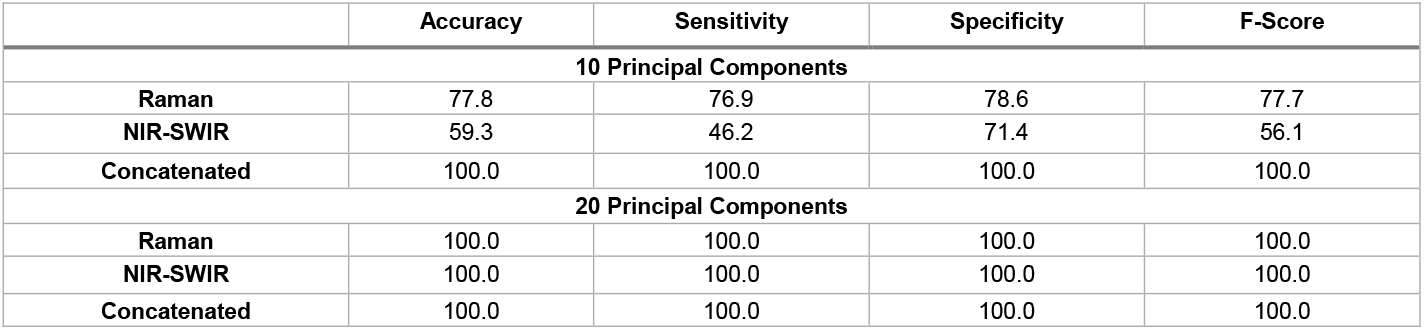
PCA-LDA Quality Parameters to quantify classification performance of PCA-LDA models built upon the foremost 10 and 20 principal components of the Raman, NIR-SWIR and Concatenated fingerprints. Ideal result is 100% for each.

|  | Accuracy | Sensitivity | Specificity | F-Score |
| --- | --- | --- | --- | --- |
| **10 Principal Components** |  |  |  |  |
| **Raman** | 77.8 | 76.9 | 78.6 | 77.7 |
| **NIR-SWIR** | 59.3 | 46.2 | 71.4 | 56.1 |
| **Concatenated** | 100.0 | 100.0 | 100.0 | 100.0 |
| **20 Principal Components** |  |  |  |  |
| **Raman** | 100.0 | 100.0 | 100.0 | 100.0 |
| **NIR-SWIR** | 100.0 | 100.0 | 100.0 | 100.0 |
| **Concatenated** | 100.0 | 100.0 | 100.0 | 100.0 |

**Fig. 2:**
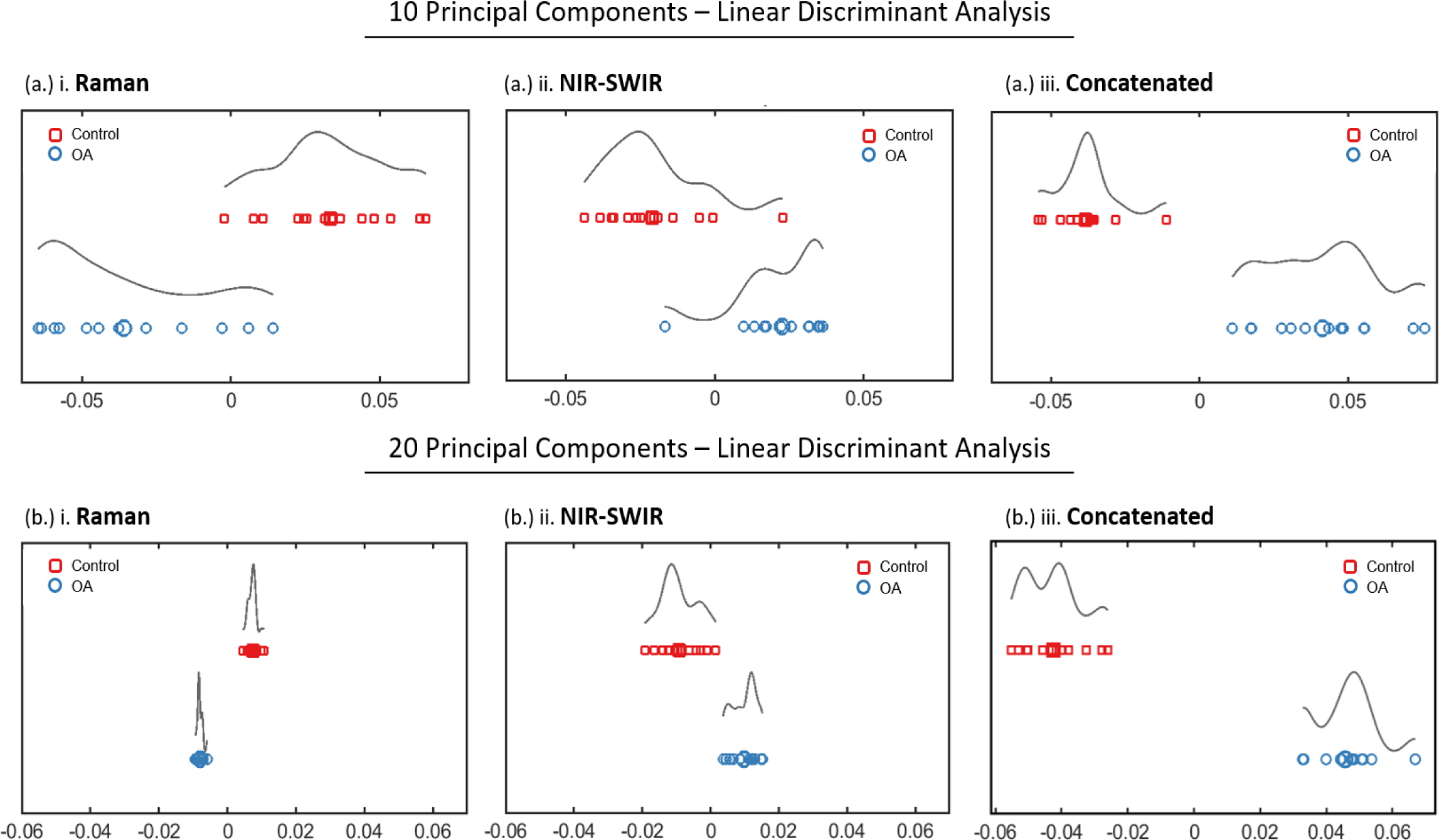
Principal Component Analysis - Linear Discriminant Analysis (PCA-LDA) classification of Raman scattering spectra, NIR-SWIR absorption scattering and the Concatenated spectra. LDA based on (a). 10 and (b). 20 principal components display marked improvement in classification for the combined spectra compared to individual analysis, illustrated by the lack of overlap and/or greater degree of separation (greater Mahanolobis distance).

Results showed a marked performance enhancement in modelling accuracy for the concatenated fingerprint over Raman and NIR-SWIR fingerprints alone. For models built on 10 PC’s accuracy increased from 77.8 % and 59.3 % for Raman and NIR-SWIR, respectively, to 100.0 % segregation of the concatenated fingerprint spectra. Models built on 20 PC’s showed classes completely segregated for all spectral modes. The difference in mean and median Mahanolobis distance were quantified as a measure of tissue classification performance. The greater the difference, the greater the discrimination between tissue classes. A marked improvement for the concatenated fingerprint over Raman and NIR-SWIR fingerprints alone was observed (summarised in Table 3).

**Table 3:** Difference between mean and median Mahalobian distance to quantify tissue classification achieved via PCA-LDA modelling of articular cartilage built upon the 20 foremost principal components of the Raman, NIR-SWIR and Concatenated fingerprints. Ideal result is a larger distance between clusters.

|  | Raman | NIR-SWIR | Concatenated |
| --- | --- | --- | --- |
| **20 PC's PCA-LDA Difference in Mahalobian distance ($\times 10^{-2}$)** |  |  |  |
| **Median** | 1.58 | 1.37 | 8.98<br>(570/ 654% increase) |
| **Mean** | 1.54 | 0.07 | 8.81<br>(572/ 1220% increase) |

An approximately 5- to 6-fold improvement in classification was demonstrated with the concatenated fingerprint over Raman and NIR-SWIR alone. These results indicate the augmented information content from both Raman and SWIR active vibrations may be responsible for improvement in tissue classification.

In the analysis using PCA and PCA-LDA, the 10 and 20 most informative components were selected from each spectral mode.^45^ A higher number of principal components described the data with greater fidelity though there was an increase in the dimensionality of the data set; a lower number may aid efficiency and speed of classification analysis.

### Machine Learning Modelling: Improved Tissue Classification under Spectromics

Spectral fingerprints formed from Raman scattering spectra, NIR-SWIR absorption spectra, and concatenated spectra were used to build a support vector machine for model “Control vs OA Cartilage” to determine tissue classification accuracy. These represented one spectrum for each of 13 osteoarthritis and 14 control model patients. Classification accuracies of SVM with 3-fold cross validation with leave-one-out cross validation shown in Fig. 3 and summarised in Table 4.

**Table 4:**
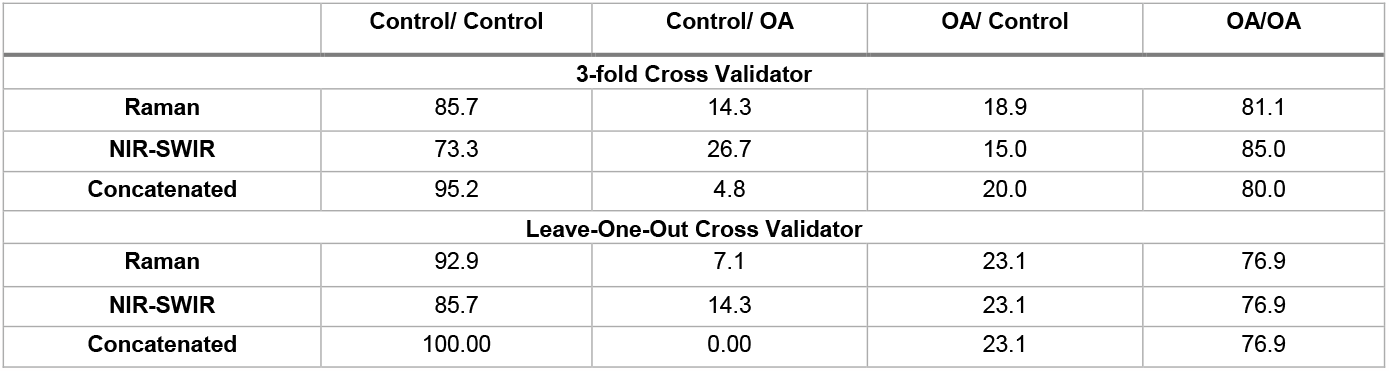
Percentage Classification Accuracies rating performance of SVM models “Control vs OA Cartilage” based on data of the Raman, NIR-SWIR and Concatenated fingerprints. Ideal result is 100% for Control/Control and OA/OA classes, and 0% for mismatched classes.

|  | Control/ Control | Control/ OA | OA/ Control | OA/OA |
| --- | --- | --- | --- | --- |
| **3-fold Cross Validator** |  |  |  |  |
| **Raman** | 85.7 | 14.3 | 18.9 | 81.1 |
| **NIR-SWIR** | 73.3 | 26.7 | 15.0 | 85.0 |
| **Concatenated** | 95.2 | 4.8 | 20.0 | 80.0 |
| **Leave-One-Out Cross Validator** |  |  |  |  |
| **Raman** | 92.9 | 7.1 | 23.1 | 76.9 |
| **NIR-SWIR** | 85.7 | 14.3 | 23.1 | 76.9 |
| **Concatenated** | 100.00 | 0.00 | 23.1 | 76.9 |

**Fig. 3:**
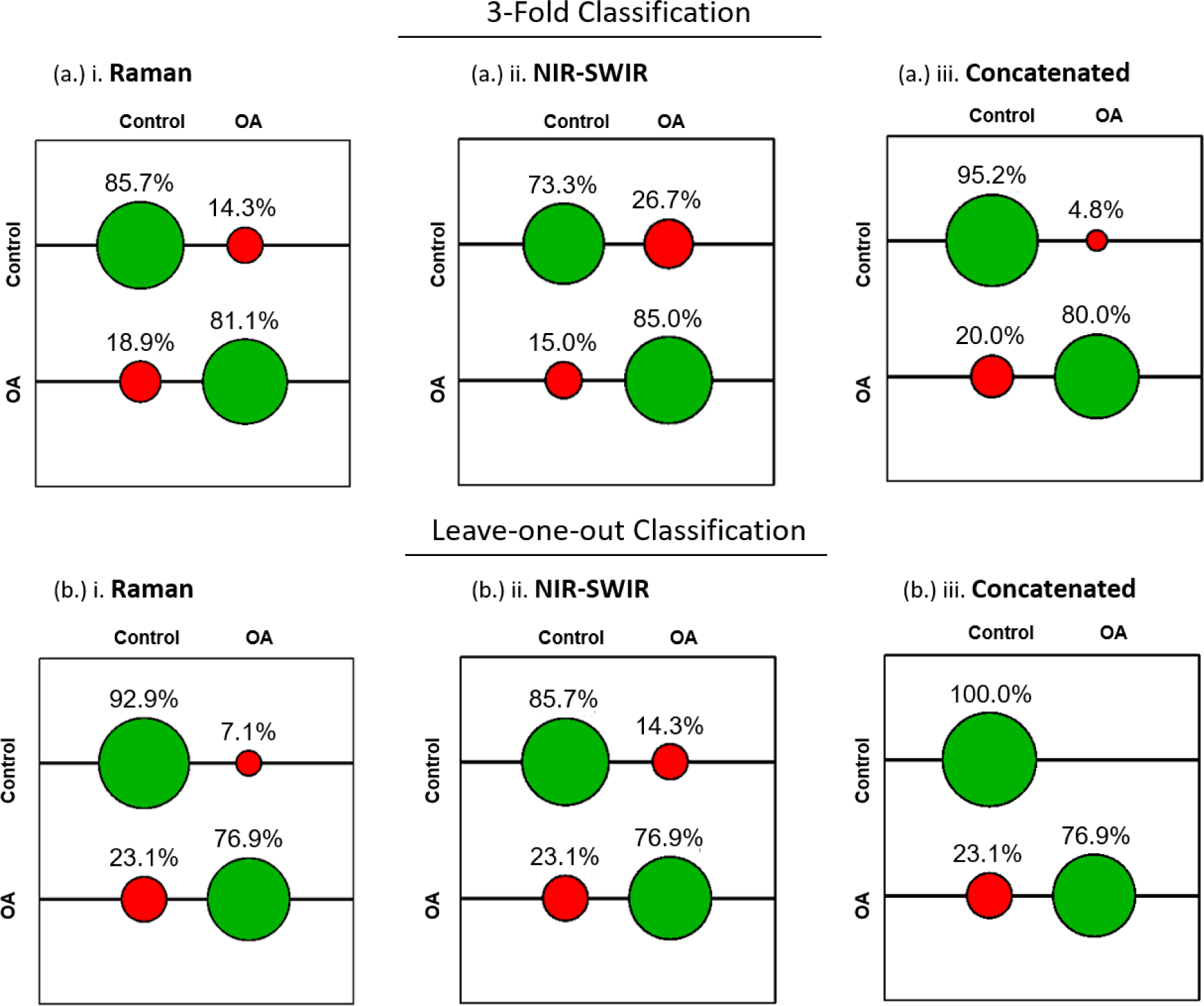
Support Vector Machine classification model (OA vs control tissue) for Raman scattering, NIR-SWIR absorption scattering and the Concatenated spectra. SVM confusion matrices (correct classification in green, incorrect in red) and associated classification rates displayed for the test dataset generated with optimal tuning parameters (c, γ) extracted from a grid search of the training dataset. The algorithm was carried out on a (a). 3-fold and (b). leave-one-out cross validator and shows improved classification when employing the concatenated spectra over Raman and NIR-SWIR alone.

The results clearly describe the improved classification under the spectromics fingerprint over modelling with Raman or NIR-SWIR alone. Specifically, recall for the control class (specificity) showed improvement from 85.7% and 73.3% for Raman and NIR-SWIR, respectively, up to 95.2% for the spectromics fingerprint under 3-fold cross validation SVM.

Under leave-one-out cross validation, where each spectrum in turn is treated as the test data and the remaining used to train the model. This approach showed an improvement again from 92.9% and 85.7%, for Raman and NIR-SWIR, respectively, to 100% accuracy for spectromics. However, recall for the positive OA class (sensitivity) showed mixed results with slightly lower rates for 3-fold SVM (Raman at 81.1%, NIR-SWIR at 85.0%, down to 80.0% for spectromics) and no difference under leave-one-out SVM observed.

Since the above evaluations were taken for each class in isolation, quality parameters were calculated from the accumulated hits to assess the classification model as a whole. Here the number of TP, TN, FP, and FN classified spectra was determined from the accumulated hits (summarised in Table 5).

**Table 5:**
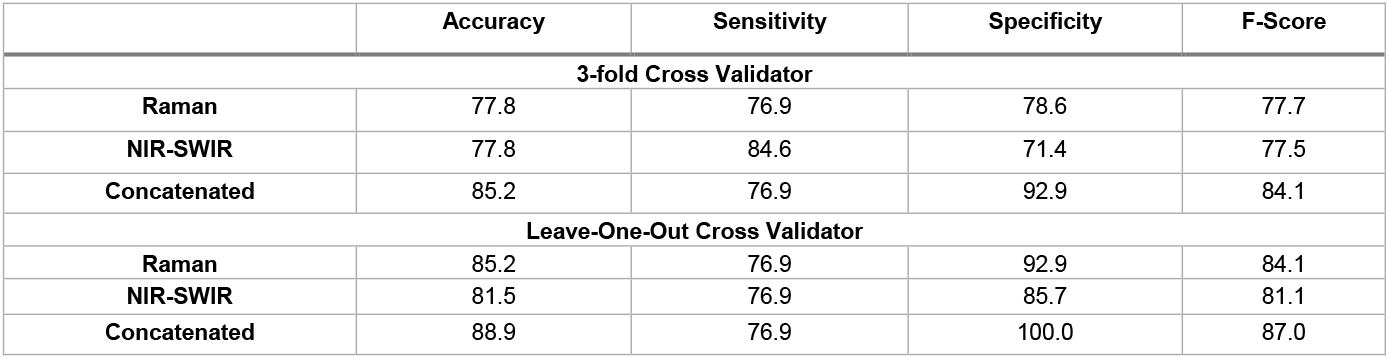
Quality Parameters based on accumulated hits of Support Vector Machine model “Control vs OA” to assess modelling performance. Ideal results are 100% for each.

|  | Accuracy | Sensitivity | Specificity | F-Score |
| --- | --- | --- | --- | --- |
| **3-fold Cross Validator** |  |  |  |  |
| **Raman** | 77.8 | 76.9 | 78.6 | 77.7 |
| **NIR-SWIR** | 77.8 | 84.6 | 71.4 | 77.5 |
| **Concatenated** | 85.2 | 76.9 | 92.9 | 84.1 |
| **Leave-One-Out Cross Validator** |  |  |  |  |
| **Raman** | 85.2 | 76.9 | 92.9 | 84.1 |
| **NIR-SWIR** | 81.5 | 76.9 | 85.7 | 81.1 |
| **Concatenated** | 88.9 | 76.9 | 100.0 | 87.0 |

Accuracy and F-Score of the whole model show marked improvement, considering the imbalanced classes, namely n=14 control and n=13 OA.^44^ Under 3-fold cross validated training, accuracy improved from 77.8% for both Raman and NIR-SWIR, to 85.2% for spectromics; F-Score from 77.7% for Raman and 77.5% for NIR-SWIR, to 84.1% for spectromics. Similar improvements were seen under leave-one-out training, with accuracy improvement from 85.2% for Raman, 81.5% for NIR-SWIR, to 88.89% for spectromics; F-score from 84.1% for Raman, 81.1% for NIR-SWIR, to 87.0% for spectromics.

Although the reported improvements were relatively modest (∼10% in specificity) this nonetheless highlights the rich chemometric information afforded by vibrational spectroscopy and the added benefit of employing the enhanced fingerprint, with specificity as high as 100%. This latter result suggests the control cartilage spectra contained greater inter-sample consistency, drawing attention perhaps to the disordering influence OA has on the tissue, resulting in relatively low sensitivity. Indeed, damage to the ECM, loss of vital PG and water, as the complex effects of the disease, would likely produce heterogeneity between patients.

Although the current data set clearly provide sufficient proof-of-concept, the work will require further validation, primarily through correlated spectromics data and an increase of number of patient samples. An increase in number of spectra will improve the training and subsequently the accuracy of both the PCA-LDA and SVM models. Thus, considering the low number of samples in each class, for which only one representative spectra (mean average) was used, the proof-of-concept demonstrates high classification performance.

### Identifying spectral regions most pertinent to OA diagnosis

To determine which regions of the fingerprint were the most significant contributors to discerning inter-class variation, a series of independent statistical tests were performed on the concatenated spectra. The results from each test (Fig. 4) were corroborated by identifying agreement between the highest scoring features (Supplementary Fig. 1). These corresponded to spectral regions most pertinent to tissue classification and in turn candidates for spectral biomarkers to distinguish control vs. OA AC tissue. These were then assigned to the chemical vibration/AC tissue constituent (Supplementary Table 1).

**Fig. 4:**
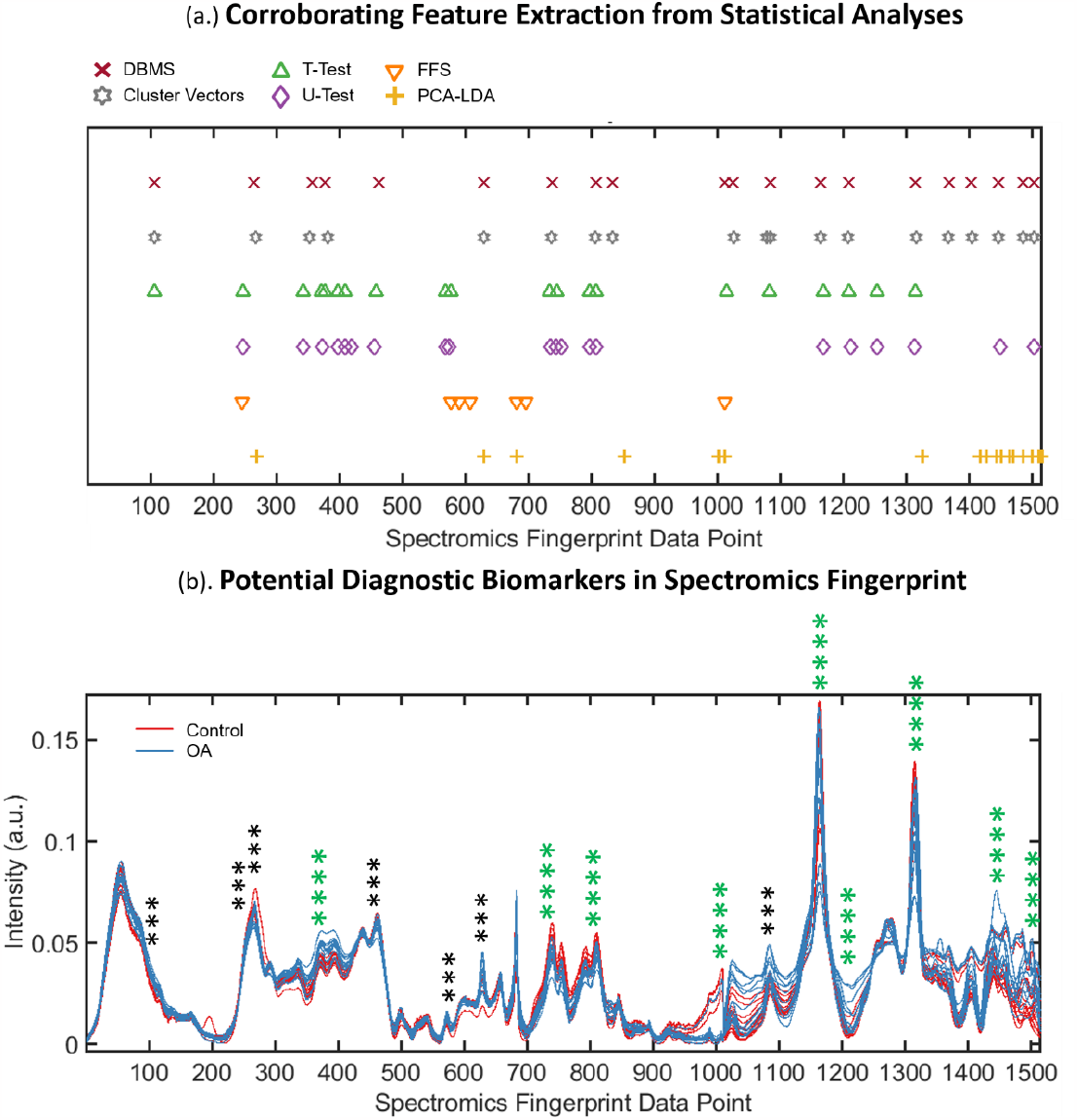
Candidate spectral biomarkers identified by corroboration between the highest scoring features extracted by separate statistical tests, (a.) Highest scoring features presented for Difference Between Mean Spectra, Cluster Vector Analysis, Student’s T-test, Mann-Whitney U-Test, Feature Forward Selection, and weightings for PCA-LDA. (b.) Corroborating the statistical tests sees certain spectral regions in agreement, displayed as regions scoring four or three hits.

Relevant Raman spectral features included peaks at 1343-1334 cm^-1^ (CH wagging) indicative of presence of glycosaminoglycan (GAG) proteins, and the typically quoted amide III band around here at 1255-1246 cm^-1^ (C-N stretching), both shown to correlate with progression of OA. ^18,25,27–29,52^ The amide II band has been employed as a prominent spectral delineator between human healthy and OA knee AC, and the red shift as an indicator of mechanical impact compression in porcine AC. ^25,31^

The peak at 1617-1616 cm^-1^ is representative of the amide I band (1612-1696 cm^-1^), with magnitude shown to decrease consistently with increasing grade of OA in human knee chondrocytes. ^53^ Spectral markers identified at 1064-1063 cm^-1^ agree strongly with the symmetric SO_3_^-^ stretching of sulphated-GAG (sGAG) protein, the quantification of which can determine the degeneration state of *ex vivo* human AC.^18,27,29^

Relevant NIR-SWIR spectral features included bands at 6543-6472 cm^-1^, attributed to the N-H stretch (-CONH, 1^st^ overtone) related to proteoglycan (PG) protein absorptions. This is regarded indicative of the ECM, and found to be the best spectral region for prediction of OA severity via Mankin scoring (6411-6496 cm^-1^). ^22^ The 4402-4366 cm^-1^ band can be attributed to the 2^nd^ overtone of the C-H bend, indicative of ECM protein content, pertinent since Partial Least Squares (PLS) modelling of the region of 4000-6000 cm^-1^ has predicted relative content of collagen and PG proteins in AC with error as low as 6%. ^25^

The features at 7159/7094 cm^-1^ and 8767/8744 cm^-1^ describe O-H stretching (1^st^ overtone) and C-H stretching (2^nd^ overtone), respectively.^18,22^ These have been attributed to water and collagen and PG proteins, their ratio forming a useful metric of relative water content (increasing during OA onset), Mankin score, and mechanical stiffness. The band centred about 5200 cm^-1^ has been used to accurately predict bound and free water through PLS regression modelling, represented by the spectromics biomarker found at 5357-5333 cm^-1. 18,19^

Interestingly, the fundamental vibrational modes of O-H and C-H stretching have been explicitly identified as Raman AC disease markers and shown to correlate with the increase in total cartilage hydration and loss of tissue constituents, respectively, as indicators of lesions. ^27^ This highlights the benefit of the complimentary and encompassing nature of the spectromics approach.

Also highlighted are potential “spectromics-specific” biomarkers, namely at the discontinuous transition from Raman to NIR-SWIR data and at around 4402-4367 cm^-1^ in NIR-SWIR absorbance. Though these cannot be attributed to pre-existing spectral markers of cartilage, these still offer a systematic feature unique to each spectra, and potentially a discriminatory metric to aid tissue classification. Further implementation of the approach could elucidate such features.

## Conclusions

This proof-of-concept study examined Raman scattering and NIR-SWIR absorption spectroscopy for chemometric assessment of articular cartilage tissue in an elegant low-level abstraction data fusion approach termed spectromics. The improved classification potential of the enhanced spectromic fingerprint has been demonstrated and the potential for diagnosis of OA.

Binary classification models based on PCA-LDA and SVM have been assessed, which exhibited notable performance improvement when built upon the holistic spectromics fingerprint over Raman or NIR-SWIR fingerprints alone. PCA-LDA segregation employing 10 PC’s improved from 77.8% and 59.3%, for Raman and NIR-SWIR, respectively, to 100% for spectromics; employing 20 PC’s showed a 5-6 fold improvement. SVM modelling similarly showed performance improvement ∼10% for class prediction using spectromics over Raman and NIR-SWIR alone. Spectral features most contributing to binary classification elucidated clinically relevant tissue components, proposed as potential label-free, non-invasive OA biomarkers.

Future work will seek to improve performance of the approach by adopting a spatially correlated spectromic fingerprinting strategy and increasing the number of patient samples. In summary, the current proof-of-concept study demonstrates the potential of the spectromics approach for OA diagnosis with significant therapeutic diagnostic implications for an aging demographic.

## Data Availability

All data produced in the present study are available upon reasonable request to the authors

## Author Contributions

*SM, RO and HC conceptualised the study. AC carried out the Raman spectroscopy work. HC acquired the NIR-SWIR spectra and developed the spectromics approach and carried out all of the analysis. DD collected the samples. KNB assisted in developing the setups for spectral acquisition. HC was supervised by both SM and RO. SM oversaw the project. HC wrote the first draft of the manuscript. SM and RO revised it. All authors contributed to the revised version and approved the final submission*.

## Acknowledgements

This work was funded by the EPSRC Doctoral Training grant (EP/N509747/1) to the School of Chemistry, University of Southampton and the EPSRC InLightenUS programme (EP/T020997/1).

## Supplementary Information

**Supplementary Figure 1:**
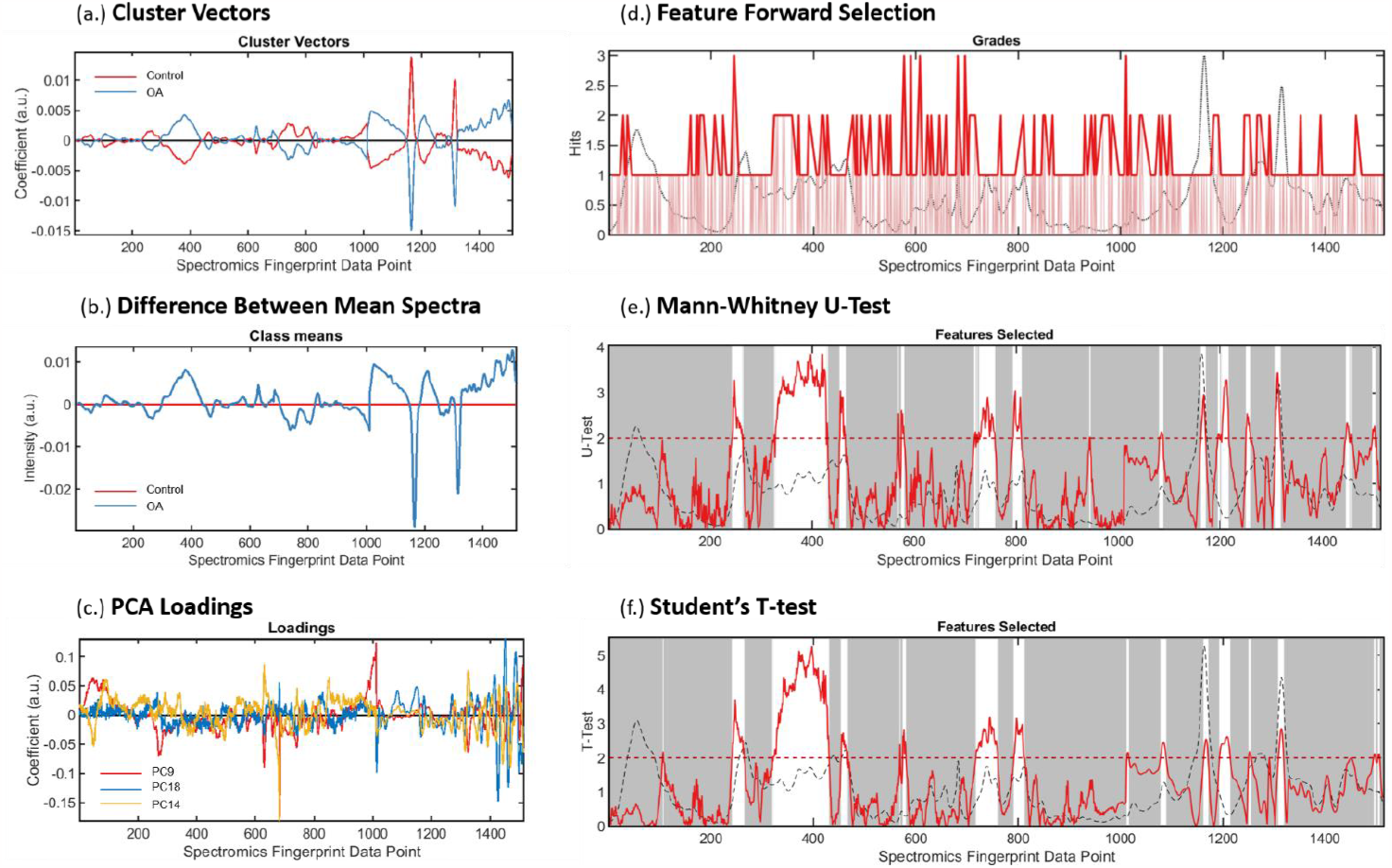
Feature selection to identify spectral biomarkers for articular cartilage diagnostics. Results were corroborated between each independent statistical analysis to highlight wavenumbers most contributing to tissue classification. (a.) Cluster Vector Analysis for class clustering, (b.) Difference Between Mean Control and OA Spectra, (c.) PCA-LDA loadings scores, (d.) Feature Forward Selection, (e.) Mann-Whitney U-Test, (f.) Student’s T-Test

**Supplementary Table 1:**
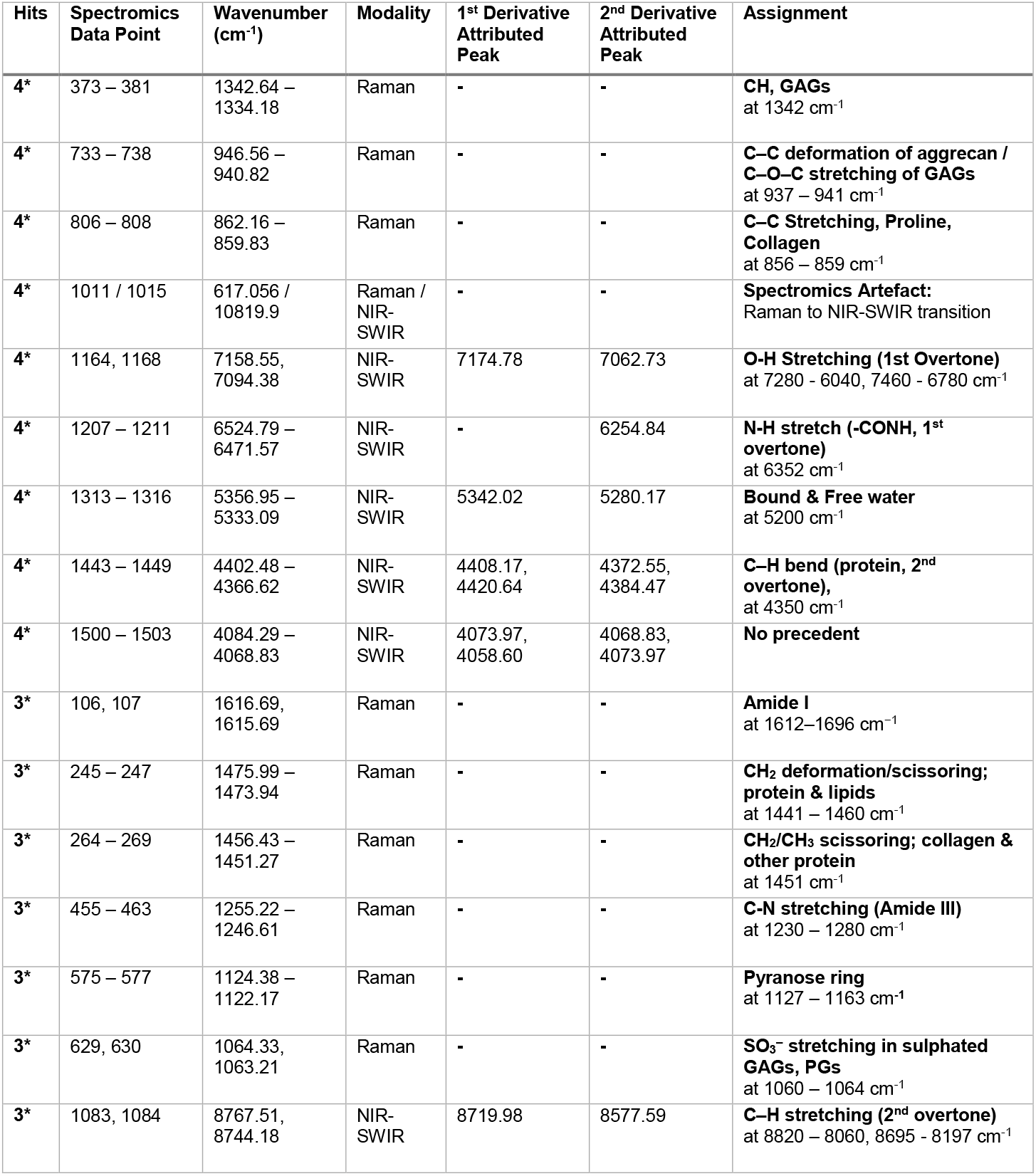
Spectral Biomarkers identified by corroboration of independent statistical tests to identify osteoarthritis vs control class discriminating peaks. Features in agreement under 3 and 4 tests (no. of hits) are displayed alongside the corresponding spectral position and attributed chemical vibration. ^18,19 18,28 53^

## Abstract Figure

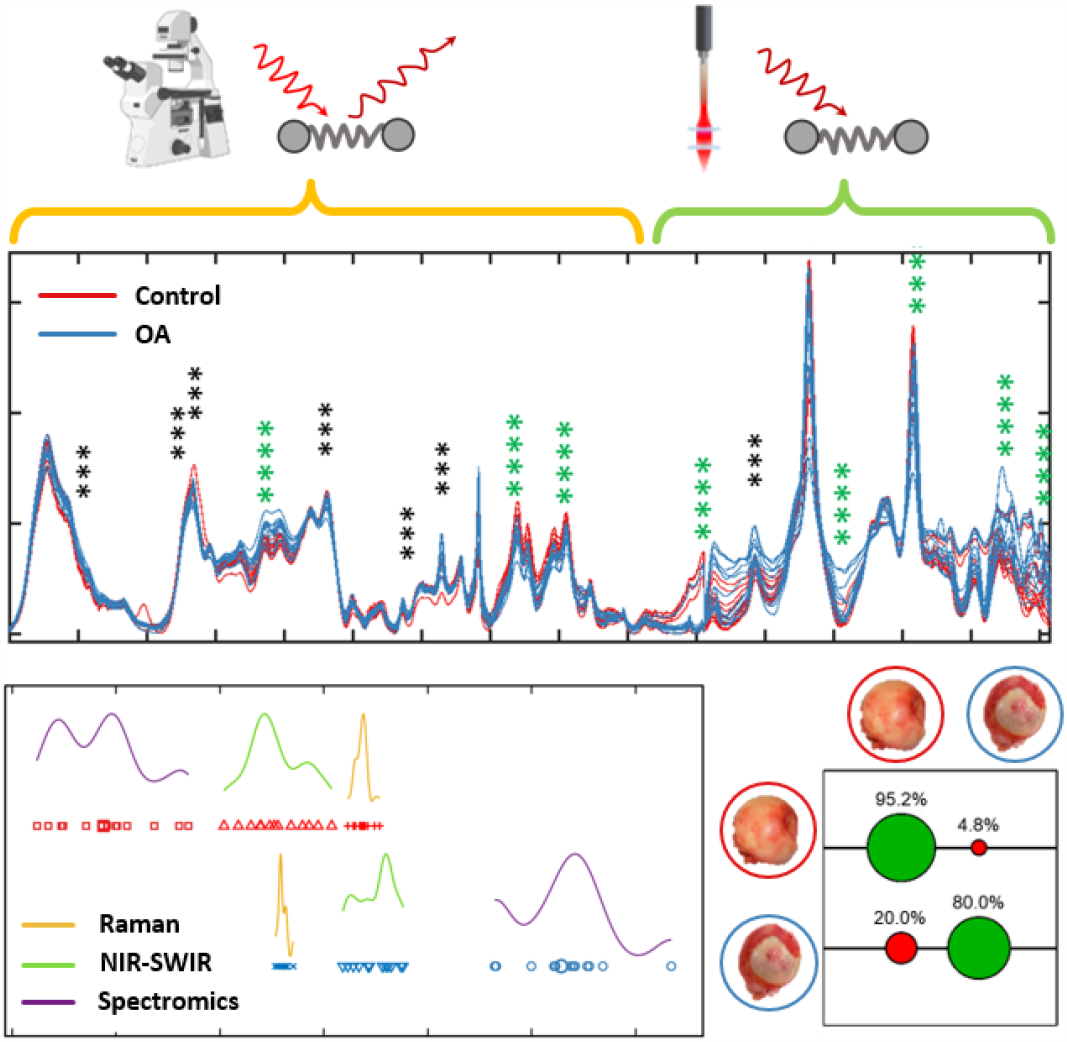

