## Supplementary Fig. 1 for "“Spectromics”: Holistic Optical Assessment of Human Cartilage via Complementary Vibrational Spectroscopy for Osteoarthritis Diagnosis"

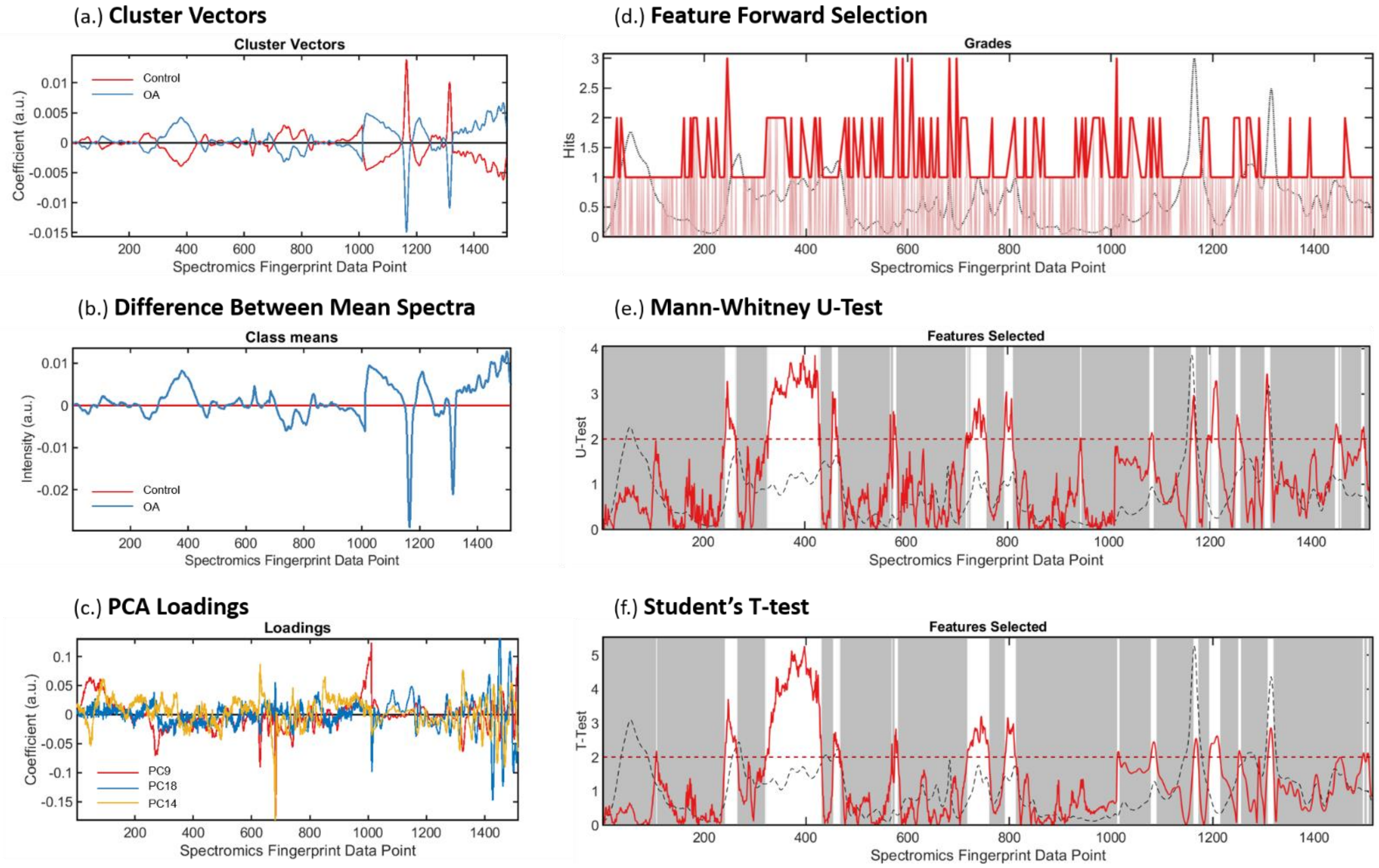


**Supplementary Figure 1:** Feature selection to identify spectral biomarkers for articular cartilage diagnostics. Results were corroborated between each independent statistical analysis to highlight wavenumbers most contributing to tissue classification. (a.) Cluster Vector Analysis for class clustering, (b.) Difference Between Mean Control and OA Spectra, (c.) PCA-LDA loadings scores, (d.) Feature Forward Selection, (e.) Mann-Whitney U-Test, (f.) Student's T-Test
