## Supplementary Table 1 for "“Spectromics”: Holistic Optical Assessment of Human Cartilage via Complementary Vibrational Spectroscopy for Osteoarthritis Diagnosis"

**Supplementary Table 1:** Spectral Biomarkers identified by corroboration of independent statistical tests to identify osteoarthritis vs control class discriminating peaks. Features in agreement under 3 and 4 tests (no. of hits) are displayed alongside the corresponding spectral position and attributed chemical vibration. [18,19] [18,28] [53]

| Hits | Spectromics Data Point | Wavenumber ($cm^{-1}$) | Modality | 1st Derivative Attributed Peak | 2nd Derivative Attributed Peak | Assignment |
|---|---|---|---|---|---|---|
| **4*** | 373 – 381 | 1342.64 – 1334.18 | Raman | **-** | **-** | **CH, GAGs** at 1342 $cm^{-1}$ |
| **4*** | 733 – 738 | 946.56 – 940.82 | Raman | **-** | **-** | **C–C deformation of aggrecan / C–O–C stretching of GAGs** at 937 – 941 $cm^{-1}$ |
| **4*** | 806 – 808 | 862.16 – 859.83 | Raman | **-** | **-** | **C–C Stretching, Proline, Collagen** at 856 – 859 $cm^{-1}$ |
| **4*** | 1011 / 1015 | 617.056 / 10819.9 | Raman / NIR-SWIR | **-** | **-** | **Spectromics Artefact:** Raman to NIR-SWIR transition |
| **4*** | 1164, 1168 | 7158.55, 7094.38 | NIR-SWIR | 7174.78 | 7062.73 | **O-H Stretching (1st Overtone)** at 7280 - 6040, 7460 - 6780 $cm^{-1}$ |
| **4*** | 1207 – 1211 | 6524.79 – 6471.57 | NIR-SWIR | **-** | 6254.84 | **N-H stretch (-CONH, 1st overtone)** at 6352 $cm^{-1}$ |
| **4*** | 1313 – 1316 | 5356.95 – 5333.09 | NIR-SWIR | 5342.02 | 5280.17 | **Bound & Free water** at 5200 $cm^{-1}$ |
| **4*** | 1443 – 1449 | 4402.48 – 4366.62 | NIR-SWIR | 4408.17, 4420.64 | 4372.55, 4384.47 | **C–H bend (protein, 2nd overtone),** at 4350 $cm^{-1}$ |
| **4*** | 1500 – 1503 | 4084.29 – 4068.83 | NIR-SWIR | 4073.97, 4058.60 | 4068.83, 4073.97 | **No precedent** |
| **3*** | 106, 107 | 1616.69, 1615.69 | Raman | **-** | **-** | **Amide I** at 1612–1696 $cm^{-1}$ |
| **3*** | 245 – 247 | 1475.99 – 1473.94 | Raman | **-** | **-** | **$CH_2$ deformation/scissoring; protein & lipids** at 1441 – 1460 $cm^{-1}$ |
| **3*** | 264 – 269 | 1456.43 – 1451.27 | Raman | **-** | **-** | **$CH_2$/$CH_3$ scissoring; collagen & other protein** at 1451 $cm^{-1}$ |
| **3*** | 455 – 463 | 1255.22 – 1246.61 | Raman | **-** | **-** | **C-N stretching (Amide III)** at 1230 – 1280 $cm^{-1}$ |
| **3*** | 575 – 577 | 1124.38 – 1122.17 | Raman | **-** | **-** | **Pyranose ring** at 1127 – 1163 $cm^{-1}$ |
| **3*** | 629, 630 | 1064.33, 1063.21 | Raman | **-** | **-** | **$SO_3^-$ stretching in sulphated GAGs, PGs** at 1060 – 1064 $cm^{-1}$ |
| **3*** | 1083, 1084 | 8767.51, 8744.18 | NIR-SWIR | 8719.98 | 8577.59 | **C–H stretching (2nd overtone)** at 8820 – 8060, 8695 - 8197 $cm^{-1}$ |
